# Associations between polyfluoroalkyl substance and organophosphate flame retardant exposures and telomere length in a cohort of women firefighters and office workers in San Francisco

**DOI:** 10.1101/2020.11.05.20226183

**Authors:** Cassidy Clarity, Jessica Trowbridge, Roy Gerona, Katherine Ona, Michael McMaster, Vincent Bessonneau, Ruthann Rudel, Heather Buren, Rachel Morello-Frosch

**Author notes:** Corresponding Author: Rachel Morello-Frosch Department of Environmental Science, Policy and Management & School of Public Health, University of California, Berkeley, 130 Mulford Hall, Berkeley CA 94702.

## Abstract

**Background:** Environmental chemical exposures can affect telomere length, which in turn has been associated with adverse health outcomes including cancer. Firefighters are occupationally exposed to many hazardous chemicals and have higher rates of certain cancers. As a potential marker of effect, we assessed associations between chemical exposures and telomere length in women firefighters and office workers from San Francisco, CA.

**Methods:** We measured serum levels of polyfluoroalkyl substances (PFAS), urinary metabolites of flame retardants, including organophosphate flame retardants (OPFRs), and telomere length in peripheral blood leukocytes in women firefighters and office workers who participated in the 2014-15 Women Workers Biomonitoring Collaborative. Multiple linear regression models were used to assess associations between chemical exposures and telomere length.

**Results:** Regression results revealed significant positive associations between perfluorooctanoic acid (PFOA) and telomere length and perfluorooctanesulfonic acid (PFOS) and telomere length among the whole cohort. Models stratified by occupation showed stronger and more significant associations among firefighters as compared to office workers. Among firefighters in models adjusted for age, we found positive associations between telomere length and log-transformed PFOA (*β*(95%CI) = 0.57(0.12, 1.02)), PFOS (0.44 (0.05, 0.83)), and perfluorodecanoic acid (PFDA) (0.43 (0.02, 0.84)). Modeling PFAS as categories of exposure showed significant associations between perfluorononanoic acid (PFNA) and telomere length among firefighters. Significant associations between OPFR metabolites and telomere length were seen for bis(1,3-dichloro-2-propyl) phosphate (BDCPP) and telomere length among office workers (0.21(0.03, 0.40)) and bis(2-chloroethyl) phosphate (BCEP) and telomere length among firefighters (−0.14(−0.28, −0.01)). For OPFRs, the difference in the direction of effect by occupational group may be due to the disparate detection frequencies and levels of exposure between the two groups and/or potential unmeasured confounding.

**Conclusion:** Our findings suggest positive associations between PFAS and telomere length in women workers, with larger effects seen among firefighters as compared to office workers. The OPFR metabolites BDCPP and BCEP are also associated with telomere length in firefighters and office workers. Associations between chemical exposures and telomere length reported here and by others suggest mechanisms by which these chemicals may affect carcinogenesis and other adverse health outcomes.

## Background

The International Agency for Research on Cancer (IARC) has designated the profession of firefighting as “possibly carcinogenic” (Group 2B)(1). Previous studies indicate that first responders and firefighters have elevated risk of various cancers including: brain, kidney, thyroid, breast, gastro-intestinal, bladder, testicular, prostate, melanoma, lymphomas, and multiple myeloma(2–10). However, most of these studies have been conducted almost exclusively on men.

There is limited research on female firefighters, despite mounting concern about breast and reproductive cancer risks among this population. This data gap is likely due to the underrepresentation of women in the fire service, however, female membership is increasing, especially in urban areas like San Francisco, which has the highest proportion of women firefighters in the US (15%)(11–13). Research on female firefighters from Daniels and colleagues showed a non-significant increase in breast cancer incidence and mortality compared to the general US population(2), while research on female firefighters in Florida has found significant increased incidence of Hodgkin’s lymphoma, thyroid cancer, cervical cancer, and brain cancer(8,14).

In addition to studying cancer, researchers have begun to examine associations between exposures to environmental chemicals and biomarkers of effect with potential relevance for cancer, including telomere length (TL)(15–19). Telomeres are complexes of repetitive DNA sequences and proteins that cap the ends of chromosomes to protect against degradation and fusion during cell division(20,21). Due to incomplete DNA replication at the terminus of DNA strands, telomeres shorten with each cell division. This attrition may be offset by the enzyme telomerase, which restores telomeric DNA(22–24). Though TL is dynamic, most cells experience net telomere shortening over the life course, eventually triggering cell senescence or apoptosis(24,25). Consequently, human TL is negatively associated with age(26–29).

Shortened telomeres have been associated with many diseases, including certain cancers(30– 36). However, telomere lengthening has also been associated with cancer(37–39), and there is evidence of an association between telomere lengthening and breast cancer(40–43). Though the exact link between TL and cancer remains unclear, research suggests mechanisms by which lengthening or shortening may contribute to carcinogenesis. For instance, telomere shortening may increase genetic instability while telomere lengthening may promote deleterious cell survival and proliferation(25,44–46).

Firefighters are occupationally exposed to many health-hazardous chemicals, including carcinogens, through activities such as fire suppression and salvage and overhaul at fire scenes(47–53). Firefighters are also exposed to hazardous chemicals in fire station dust, diesel exhaust, firefighting foams, contaminated fire equipment, and certain firefighting gear(54–59). Studies have documented firefighters’ exposure to benzene, polycyclic aromatic hydrocarbons (PAHs), formaldehyde, dioxins, polybrominated diphenyl ethers (PBDEs), polyfluoroalkyl substances (PFAS), and organophosphate flame retardants (OPFRs)(60–68).

PFAS, which are widely used for their ability to impart grease, stain, and water resistance to items such as food packaging, non-stick cookware, paints, fabrics, carpets, and furniture(69), are of particular concern for firefighters. Firefighters are exposed to PFAS through the combustion of PFAS-containing products such as furniture and carpet and through firefighting gear and firefighting foams that contain these compounds(57,64,70,71). Indeed, research shows that firefighters have elevated levels of certain PFAS relative to non-firefighters(64,67,71). PFAS exposures have been associated with adverse health outcomes, including cancer(72–79).

Firefighters are also occupationally exposed to flame retardants(65,68,80). While use of polybrominated diphenyl ether (PBDE) flame retardants in consumer products has been gradually phased out due to their toxicity to humans, persistence in the environment, and ability to bioaccumulate(81), OPFRs and other halogenated flame retardants have emerged as replacements and have been found in fire stations(82–84). There are few epidemiological studies on the human health effects of OPFRs. The existing literature shows associations between OPFR levels in house dust and urine and decreased sperm quality and hormone dysfunction in men (85), and lower thyroxine levels in women with higher urinary levels of diphenyl phosphate(86). In experimental studies, OPFRs cause endocrine disruptions in sex hormones and thyroid hormones(75,87–91).

Experimental studies on chemical exposures and telomere length are limited and findings are inconsistent(92). Previous epidemiological studies have found PFAS exposures are associated with altered TL in humans; perfluorooctanoic acid (PFOA) has been associated with telomere shortening(18) and perfluorooctane sulfonic acid (PFOS) has been associated with telomere lengthening(17). To our knowledge, no human studies have examined associations between OPFRs and TL, although one study of chemically similar organophosphate insecticides found associations with altered TL, with the direction of effect depending on the insecticide in question(93). As a possible intermediary between exposure and disease, TL serves as a biomarker of effect for assessing the potential impacts of environmental exposures on human health.

To better characterize firefighters’ exposures with relevance to women’s health outcomes, a collaboration of firefighters, scientists, and environmental health advocates created a community-based participatory research project, the Women Workers Biomonitoring Collaborative (WWBC). We interviewed female firefighters and office workers in San Francisco, CA, and collected biospecimens (urine and serum). Samples were analyzed for PFAS (serum), flame retardant metabolites (urine), and telomere length (whole blood leukocytes). We then assessed the relationship between PFAS and flame retardant metabolite levels and TL in female firefighters and office workers.

## Methods

### Recruitment and consent

The WWBC recruitment, enrollment, and sample collection protocol has been described previously(67). Briefly, recruitment and sample collection took place between June 2014 and March 2015. Firefighter study partners from the San Francisco Fire Department (SFFD) and researchers collaborated on recruitment of both firefighter from SFFD and office worker participants from the City and County of San Francisco. Study inclusion criteria included self-identifying as female, being over 18 years of age, full-time employment, and being a nonsmoker. Additionally, firefighters were required to have at least 5 years of service with the SFFD and to be on “active duty” (i.e., assigned to a fire station) at the time of recruitment. Informed consent was obtained from all participants prior to data collection activities following protocols approved by the Institutional Review Board of the University of California, Berkeley (#2013-07-5512).

### Data collection and sample processing

Each participant completed an hour-long exposure assessment interview that captured demographics, basic health information, and possible sources of chemical exposure from occupational activities, consumer product use, and diet. A certified phlebotomist collected blood samples in 10 mL additive-free glass tubes and 10 mL EDTA glass tubes. Urine was collected in 60 mL polypropylene specimen cups. All samples were transported in a cooler with ice and processed within 3 hours of collection. The serum was separated by allowing clotting at room temperature followed by centrifuging at 3000 rpm for 10 minutes. The serum and whole blood were aliquoted into 1.2 mL cryovials and urine into 3.5 mL cryovials and stored at −80 °C until analysis. All samples were processed and analyzed at the University of California, San Francisco.

### PFAS analysis

As described previously(67), twelve PFAS were selected for targeted analysis in serum: perfluorobutanoic acid (PFBA), perfluorohexanoic acid (PFHxA), perfluoroheptanoic acid (PFHpA), perfluorooctanoic acid (PFOA), perfluorononanoic acid (PFNA), perfluorodecanoic acid (PFDA), perfluoroundecanoic acid (PFUnDA), perfluorododecanoic acid (PFDoA), perfluorobutane sulfonic acid (PFBuS), perfluorohexane sulfonic acid (PFHxS), perfluorooctane sulfonic acid (PFOS), and perfluorooctane sulfonamide (PFOSA). The 12 PFAS were analyzed in 0.5 mL of serum using liquid chromatography-tandem mass spectrometry (LC-MS/MS). An Agilent LC1260 (Sta. Clara, CA)-AB Sciex API 5500 (Foster City, CA) platform was used in the analysis. Each sample was prepared for analysis by solid phase extraction using a Waters Oasis HLB cartridge (10 mg, 1cc). Extracted aliquots of each sample (25 uL) were run in duplicates. The 12 analytes were separated by elution gradient chromatography using Phenomenex Kinetex C18 column (100 x 4.6 mm, 2.6 µ) at 40°C. Electrospray ionization (negative mode) was used as method of ionization for individual analytes.

Analytes were detected in each sample by multiple reaction monitoring using two transitions per analyte. To determine the presence of each analyte, retention time matching (within 0.15 min) along with the peak area ratio between its qualifier and quantifier ions (within 20%) were used. Quantification of each detected analyte was done by isotope dilution method using a 10-point calibration curve (0.02-50 ng/mL) and employing two C13-labelled PFAS isotopologues. Procedural quality control materials and procedural blanks were run along with the calibration curve at the start, middle, and end of each run. Two QC materials were used at low and high concentrations. To accept the results of a batch run, QC materials measurements must be within 20% of their target values and the precision of their measurements within 20% CV (coefficient of variation).

The limits of quantification for the 12 analytes range from 0.05 to 0.1 ng/mL. Analyte identification from total ion chromatograms was evaluated using AB Sciex Analyst v2.1 software while quantification of each analyte was processed using AB Sciex MultiQuant v2.02 software. Analysts were blinded to firefighter and office worker status of the serum samples during the analysis. Results were reported in ng/mL for all 170 study participants(67).

### Flame retardant analysis

We quantified metabolites of six OPFR chemicals in urine: bis(1,3-dichloro-2-propyl) phosphate (BDCPP), bis(2-chloroethyl) phosphate (BCEP), dibutyl phosphate (DBuP), dibenzyl phosphate (DBzP), di-p-cresyl phosphate (DpCP), di-o-cresyl phosphate (DoCP), and 4 brominated flame retardants: 2,3,4,5-tetrabromobenzoic acid (TBBA), tetrabromobisphenol a (TBBPA), 5-OH-BDE 47, and 5-OH-BDE 100.

Quantitative analysis was performed using liquid-chromatography-tandem mass spectrometry (LC-MS/MS) on an Agilent LC 1260 (Agilent Technologies, Sta. Clara, CA)-AB Sciex 5500 system (Sciex, Redwood City, CA). Freshly thawed urine specimens (1 mL) were deconjugated prior to LC-MS/MS analysis by addition of 450 U H. pomatia glucuronidase (Sigma-Aldrich, St Louis, MO) and incubated at 37 °C for two hours with constant shaking. Deconjugated urine samples were prepared for LC-MS/MS analysis by solid phase extraction (SPE) using Waters Oasis WAX cartridges (10 mg, 30 μm, 1 cc). Analytes in the extracted aliquots were separated by elution gradient chromatography using an Agilent ZORBAX Eclipse XDB-C8 column (2.1×100 mm, 3.5um) maintained at 50°C. Negative mode electrospray ionization (ESI) was used to ionize analytes and mass scanning was performed by multiple reaction monitoring. Each analyte was monitored using two transitions and retention time. Quantitation of each analyte was performed by isotope dilution method with their deuterated or C-13 isotopologues as internal standards.

Each sample was injected in duplicate. Procedural quality control materials and procedural blanks were run along with the calibration curve at the start, middle, and end of each run. Two QC materials were used at low and high concentrations. To accept the results of a batch run, QC materials measurements must be within 20% of their target values and the precision of their measurements have ≤ 20% CV (coefficient of variation). Analyte identification from total ion chromatograms was evaluated using AB Sciex Analyst v2.1 software while quantification of each analyte was processed using AB Sciex MultiQuant v2.02 software. Analysts were blinded to firefighter and office worker status of the urine samples during the analysis.

### Telomere analysis

DNA was extracted from 200 µL of whole blood using the Qiagen Qamp Mini Blood Kit (cat. No. 51104) according to the manufacturer’s instructions. One microgram of DNA from the samples was digested with Hinfl and RsaI, run on 0.8% TAE gels for 6 hours and Southern transferred to Nylon membranes. The membranes were hybridized with digoxigenin-labeled telomere probes (Sigma TeloTAGGG Telomere Length Assay, cat. No. 12209136001) followed by incubation with anti-digoxigenin alkaline phosphatase conjugates. DNA bands were detected using chemilunescence and analyzed using ImageQuant software (GE Healthcare). The mean terminal restriction fragment (TRF) length was derived from standards provided in the kit. Results were reported in kilobase pairs (kbp) for 163 participants.

### Statistical analysis

All chemical distributions were skewed and log-transformed using natural logarithms to improve normality. Descriptive statistics such as geometric mean (GM), geometric standard deviation (GSD), and 95% confidence intervals (CI) were calculated for TL, PFAS, and flame retardants with ≥ 60% detection frequency in at least one occupational group.

To assess the relationship between chemical exposure and TL, we developed linear regression models for each compound. Chemicals with ≥ 70% detection frequency were included in linear models as continuous predictor variables. For models using continuous PFAS levels as a predictor, values below the limit of detection (LOD) were substituted with LOD/√2. For models using continuous flame retardant metabolite levels as a predictor, we included all LC-MS/MS reported values, including those reported below the LOD, and substituted LOD/√2 for any remaining non-detected values.

For chemicals with < 70% detection frequency but ≥ 40% detection frequency, chemical concentrations were categorized as either <LOD, LOD-50^th^%, and >50^th^% or as <LOD, associations between perfluorononanoic2265;LOD, depending on detection frequencies. Telomere data were roughly normally distributed and were therefore not log-transformed.

Potential confounders were selected *a priori* based on results from previous literature and prior analyses performed on this data(67). Covariates assessed include demographic variables such as race/ethnicity and education; health variables such as body mass index (BMI), stress, and sleep metrics; and food frequency variables. Spearman correlations were used to test independent relationships between continuous covariates and telomere length and chemical predictors using the Benjamini-Hochberg procedure to control for multiple testing(94). Analysis of Variance (ANOVA) and t-tests or Wilcoxon rank sum tests were performed to assess differences in TL or chemical predictors across categorical and dichotomous covariates, respectively. Covariates were included in final models as confounders if they had a significant association (p≤0.10) with both TL (the outcome) and at least one chemical congener (the exposure). For PFAS models, ANOVA for nested models and assessment of percent change in coefficients (Δ≥10%) were also used to inform variable selection.

As exploratory analyses revealed disparate effect estimates by occupation for both PFAS and flame retardant metabolites, models were run on the entire study population where possible and also stratified by occupation. The following equation was used to interpret results from models with a continuous log-transformed chemical predictor: (*β* ∗ *ln*(1 + (*x*/100)∗ 1000, where x equals the percent change in PFAS exposure and multiplying by 1000 provides a result in base pairs as opposed to kilobase pairs. For categorical models, raw model estimates in kilobase pairs were multiplied by 1000 for results in base pairs. All analyses were performed in R version 3.5.1 and R studio version 1.1.463(95,96).

### PFAS models

We developed minimally adjusted models (Model 1) and fully adjusted models (Model 2) to assess the association between continuous log-transformed PFAS (logPFAS) and TL. Due to the well-documented correlation between age and TL(26–29), age was included in all models. Model 1 was adjusted for continuous age in years. Based on covariate tests described above, Model 2 was adjusted for age, occupation, the number of times dairy products were eaten per week, and the number of times eggs were eaten per week. Models were run for the full cohort and stratified by occupation.

Data visualization using locally weighted regression (loess) curves mapped onto bivariate scatter plots of TL and logPFAS suggested potential non-linear relationships that compelled us to run PFAS Model 2 with PFHxS, PFOA, PFOS, PFNA, and PFDA categorized into quartiles. PFUnDA and PFBuS were categorized into tertiles due to low detection frequency.

### Flame retardant models

Few covariates were associated with flame retardant metabolites within this population(68). Age was included in all models as well as log-transformed creatinine (logCreatinine) to account for differences in urine dilution(97).

Only BDCPP had sufficient detection frequency to model as a continuous variable for both firefighters and office workers. To test if effect estimates of the BDCPP-TL relationship varied significantly by occupation, we added an interaction term to the full BDCPP-TL model.

Among firefighters, BCEP and DBuP had sufficient detection frequency to include in models as continuous variables though these metabolites were also modeled as categorical variables for comparison across occupations. TBBPA and DpCP were analyzed as categorical variables. The firefighter data for BCEP and DBuP were categorized as <LOD, LOD-50^th^%, and >50^th^% since the detection frequencies were greater than 50%, which allowed for categorization into three groups. Office worker data for BCEP and DbuP and all TBBPA and DpCP data were categorized as <LOD and ≥ LOD due to detection frequencies below 50%. Models for these categorized compounds were stratified by occupation.

## Results

In total, 176 participants enrolled in the study. Six participants (three firefighters and three office workers) were dis-enrolled or did not provide biospecimen samples, and seven participants (two firefighters and five office workers) did not have adequate sample to perform the telomere analysis. The final study sample consisted of 84 firefighters and 79 office workers (N = 163) (**Table 1**). Firefighters had longer telomeres than office workers, but were otherwise similar to office workers in age, dairy consumption, and egg consumption. A detailed description of firefighter and office worker differences in the WWBC has been previously reported(67). In brief, office workers were more often born outside the US, married, worked at the City and County of San Francisco for less time, and had higher educational attainment levels compared to firefighters, although firefighters had higher incomes. Race/ethnicity and BMI were similar across groups(67). These variables were not associated with TL in our population.

**Table 1.** Descriptive statistics for model parameters by occupation.

|  | Full cohort<br>(N=163)<br>Mean(SD) or<br>GM(GSD)<sup>a</sup> | Firefighters<br>(N=84)<br>Mean(SD) or<br>GM(GSD)<sup>a</sup> | Office Workers<br>(N=79)<br>Mean(SD) or<br>GM(GSD)<sup>a</sup> | p-value<sup>b</sup> | LOD<sup>c</sup> | Full<br>cohort<br>DF<sup>d</sup>(%) | Fire-<br>fighters<br>DF(%) | Office<br>workers<br>DF(%) |
| --- | --- | --- | --- | --- | --- | --- | --- | --- |
| **Age (years)** |  |  |  |  |  |  |  |  |
|  | 47.93(8.10) | 47.38(4.58) | 48.52(10.64) | 0.23 | - | - | - | - |
| **Dairy consumption (times per week)** |  |  |  |  |  |  |  |  |
|  | 14.45(8.19) | 13.98(7.30) | 14.96(9.07) | 0.47 | - | - | - | - |
| **Egg consumption (times per week)** |  |  |  |  |  |  |  |  |
|  | 3.71(1.93) | 3.96(2.00) | 3.46(1.84) | 0.23 | - | - | - | - |
| **Telomere Length (mean TRF in kbp<sup>e</sup>)** |  |  |  |  |  |  |  |  |
|  | 7.90(1.12) | 8.10(1.09) | 7.68(1.13) | 0.01* | - | - | - | - |
| **PFAS (ng/mL)** |  |  |  |  |  |  |  |  |
| PFHxS | 3.68(2.79) | 4.55(2.82) | 2.94(2.64) | 0.01* | 0.02 | 100 | 100 | 100 |
| PFOA | 1.16(1.76) | 1.13(1.70) | 1.19(1.83) | 0.44 | 0.02 | 100 | 100 | 100 |
| PFOS | 4.18(2.08) | 4.33(1.83) | 4.03(2.35) | 0.39 | 0.02 | 100 | 100 | 100 |
| PFNA | 0.69(1.94) | 0.77(1.98) | 0.61(1.87) | 0.09 | 0.05 | 100 | 100 | 100 |
| PFDA | 0.26(2.09) | 0.27(1.77) | 0.24(2.42) | 0.31 | 0.02 | 99 | 100 | 98 |
| PFUnDA | 0.18(4.44) | 0.23(3.67) | 0.14(5.19) | 0.06 | 0.02 | 80 | 87 | 73 |
| PFBuS | 0.13(4.28) | 0.13(4.19) | 0.13(4.42) | 0.85 | 0.02 | 73 | 74 | 72 |
| **OPFRs (ng/mL)** |  |  |  |  |  |  |  |  |
| BDCPP | 1.92(5.11) | 4.05(4.59) | 0.87(3.85) | ≤0.01* | 0.20 | 95 | 100 | 90 |
| BCEP | 0.40(6.11) | 0.87(5.72) | --<sup>f</sup> | --<sup>g</sup> | 0.10 | 60 | 79 | 39 |
| DBuP | --<sup>f</sup> | 0.41(3.95) | --<sup>f</sup> | --<sup>g</sup> | 0.10 | 56 | 82 | 28 |
| TBBPA | --<sup>f</sup> | --<sup>f</sup> | --<sup>f</sup> | --<sup>g</sup> | 0.20 | 44 | 46 | 41 |
| DpCP | --<sup>f</sup> | --<sup>f</sup> | --<sup>f</sup> | --<sup>g</sup> | 0.10 | 29 | 42 | 16 |

<sup>a</sup> Geometric mean (GM) and geometric standard deviation (GSD) computed for egg consumption, all PFAS, and all OPFRs due to skewed distributions
<sup>b</sup> p-values derived from Wilcoxon rank-sum tests
<sup>c</sup> LOD = limit of detection
<sup>d</sup> DF = detection frequency
<sup>e</sup> kilobase pairs
<sup>f</sup> GMs were not calculated for chemicals with detection frequencies below 60%
<sup>g</sup> Wilcoxon rank-sum tests were not performed for chemicals with detection frequencies below 60% in any group
<sup>f</sup> All summary statistics calculated using LOD/sqrt(2) for those chemicals with less than 100% detection frequency

### PFAS exposure and telomere length

We measured serum for 12 PFAS, four of which (PFBA, PFHxA, PFHpA, and PFOSA) had no measurable levels above the LOD in any participant. Seven PFAS congeners had detection frequencies greater than 70%, of which four had detection frequencies of 100% (PFHxS, PFOA, PFOS, PFNA). PFHxS was found at significantly higher levels among firefighters compared to office workers. Higher levels of PFNA were also observed among firefighters, however the group difference was not statistically significant in this subset of WWBC data. Distributions of the remaining PFAS were similar across groups (**Table 1**). A full description of differences and predictors of PFAS levels in firefighters and office workers is described elsewhere(67).

Of the covariates assessed as potential confounders of the PFAS-TL relationship, only age, occupation, dairy consumption, and egg consumption met our criteria for inclusion in fully adjusted models. Effect estimates were generally larger among firefighters compared to office workers (**Table 2 and Additional file 1**). In both models, exposure to PFOA and PFOS was associated with significantly longer TL among the entire cohort. In Model 1, a doubling (or 100 percent increase) of PFOA concentration was associated with a 273 (95% CI 54, 493) base pair (bp) increase in TL. In Model 2, a doubling in PFOA was associated with a 240 (95% CI 25, 455) bp increase in TL. A doubling in PFOS concentration was associated with a 183 (95% CI 15, 352) bp increase in TL in Model 1, and a 172 (95% CI 5, 340) bp increase in TL in Model 2 (**Table 2**).

**Table 2.**
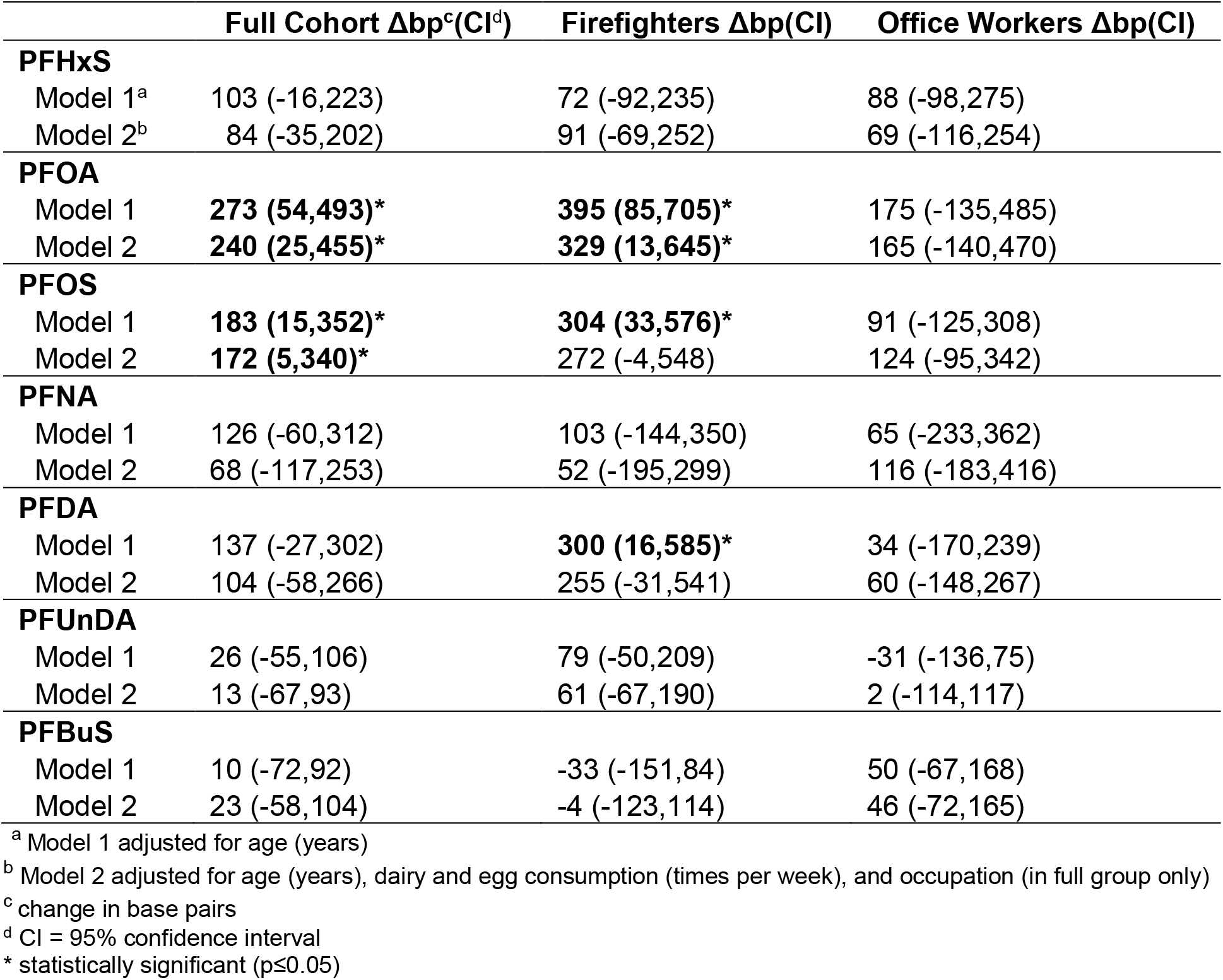
Estimated base pair change in telomere length for a doubling of PFAS concentration.

| | Full Cohort $\Delta bp^c(CI^d)$ | Firefighters $\Delta bp(CI)$ | Office Workers $\Delta bp(CI)$ |
| --- | --- | --- | --- |
| **PFHxS** |  |  |  |
| Model 1<sup>a</sup> | 103 (-16,223) | 72 (-92,235) | 88 (-98,275) |
| Model 2<sup>b</sup> | 84 (-35,202) | 91 (-69,252) | 69 (-116,254) |
| **PFOA** |  |  |  |
| Model 1 | **273 (54,493)\*** | **395 (85,705)\*** | 175 (-135,485) |
| Model 2 | **240 (25,455)\*** | **329 (13,645)\*** | 165 (-140,470) |
| **PFOS** |  |  |  |
| Model 1 | **183 (15,352)\*** | **304 (33,576)\*** | 91 (-125,308) |
| Model 2 | **172 (5,340)\*** | 272 (-4,548) | 124 (-95,342) |
| **PFNA** |  |  |  |
| Model 1 | 126 (-60,312) | 103 (-144,350) | 65 (-233,362) |
| Model 2 | 68 (-117,253) | 52 (-195,299) | 116 (-183,416) |
| **PFDA** |  |  |  |
| Model 1 | 137 (-27,302) | **300 (16,585)\*** | 34 (-170,239) |
| Model 2 | 104 (-58,266) | 255 (-31,541) | 60 (-148,267) |
| **PFUnDA** |  |  |  |
| Model 1 | 26 (-55,106) | 79 (-50,209) | -31 (-136,75) |
| Model 2 | 13 (-67,93) | 61 (-67,190) | 2 (-114,117) |
| **PFBuS** |  |  |  |
| Model 1 | 10 (-72,92) | -33 (-151,84) | 50 (-67,168) |
| Model 2 | 23 (-58,104) | -4 (-123,114) | 46 (-72,165) |

<sup>a</sup> Model 1 adjusted for age (years)
<sup>b</sup> Model 2 adjusted for age (years), dairy and egg consumption (times per week), and occupation (in full group only)
<sup>c</sup> change in base pairs
<sup>d</sup> CI = 95% confidence interval
\* statistically significant ( $p \leq 0.05$ )

Among firefighters, exposure to PFOA, PFOS, and PFDA was significantly associated with longer TL in Model 1. In Model 2 (adjusted for age, dairy consumption, and egg consumption), only PFOA remained significantly associated with TL. Among firefighters, a doubling of PFOA concentration was associated with a 395 (95% CI 85,705) base pair (bp) increase in TL in Model 1, and a 329 (95% CI 13,645) bp increase in TL in Model 2. In Model 1, a doubling in firefighters’ PFOS concentration was associated with a 304 (95% CI 33,576) bp increase in TL, and a doubling in firefighters’ PFDA concentration is associated with a 300 (95% CI 16,585) bp increase in TL.

Most PFAS were positively associated with TL in office workers, though effect estimates were smaller than for firefighters and none were statistically significant. No interaction terms testing for effect modification by occupation were statistically significant.

To assess the shape of the exposure-response relationships, we modeled locally weighted regression (loess) curves atop unadjusted scatter plots of TL and logPFAS, stratified by occupation. Among firefighters, the loess curves suggested potential non-linear exposure-response relationships, with log-transformed PFOA, PFOS, PFNA, and PFDA exhibiting a somewhat conserved pattern (**Figure 1**). In firefighters, exposure to these four PFAS compounds appears to be associated with increasing TL from low to intermediate concentrations and unchanging or decreasing TL at higher concentrations.

**Figure 1.**
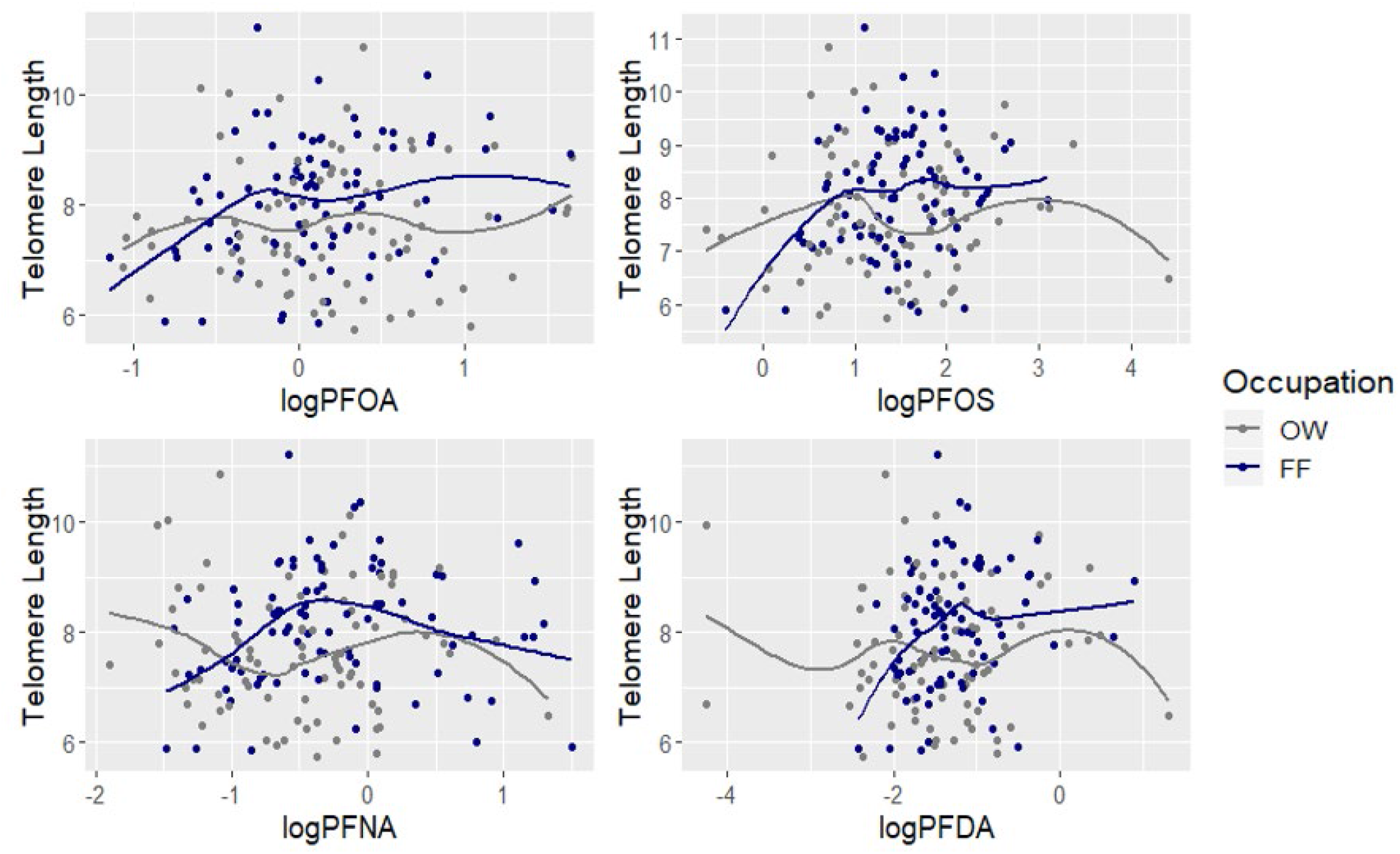
Loess curves^a^ for telomere length and log-transformed PFAS by occupation. ^a^ Locally-weighted regression with span = 0.75

To further explore this relationship, we included PFAS as categorical variables in fully adjusted models. **Table 3** details the estimated base pair change for a categorical increase of PFAS exposure relative to the referent (first quartile) from fully adjusted models. Among firefighters, PFNA, which had non-statistically significant associations with TL in the continuous linear Models 1 and 2, had significant effect estimates for each quartile of exposure relative to the referent, with the greatest increase in the second quartile as was suggested by the loess curve.

**Table 3.** Estimated^a^ base pair change in telomere length relative to the reference group.

| | Firefighters $\Delta bp^b(CI^c)$ | Office Workers $\Delta bp(CI)$ |
| --- | --- | --- |
| **PFHxS** |  |  |
| Reference<sup>d</sup> | - | - |
| Q2 | -62 (-728, 604) | 209 (-515, 933) |
| Q3 | -180 (-850, 491) | 238 (-522, 998) |
| Q4 | 366 (-301, 1033) | 163 (-563, 889) |
| **PFOA** |  |  |
| Reference | - | - |
| Q2 | 209 (-464, 882) | 10 (-704, 724) |
| Q3 | 290 (-387, 967) | 500 (-197, 1198) |
| Q4 | 368 (-327, 1063) | 143 (-584, 869) |
| **PFOS** |  |  |
| Reference | - | - |
| Q2 | 211 (-453, 876) | 301 (-417, 1019) |
| Q3 | 599 (-82, 1280) | -18 (-716, 681) |
| Q4 | 397 (-288, 1082) | 426 (-357, 1210) |
| **PFNA** |  |  |
| Reference | - | - |
| Q2 | **1250 (631, 1869)\*** | -344 (-1064, 376) |
| Q3 | **809 (197, 1421)\*** | -46 (-769, 676) |
| Q4 | **668 (29, 1307)\*** | 164 (-576, 904) |
| **PFDA** |  |  |
| Reference | - | - |
| Q2 | 187 (-472, 845) | -4 (-729, 721) |
| Q3 | 614 (-48, 1275) | -6 (-722, 710) |
| Q4 | 396 (-265, 1058) | 245 (-489, 978) |
| **PFUnDA** |  |  |
| Reference | - | - |
| T2 | 22 (-549, 592) | -154 (-774, 466) |
| T3 | 488 (-88, 1065) | 215 (-430, 861) |
| **PFBuS** |  |  |
| Reference | - | - |
| T2 | -184 (-782, 414) | 144 (-503, 791) |
| T3 | -19 (-606, 569) | 15 (-598, 628) |

<sup>a</sup> Model adjusted for age (years) and dairy and egg consumption (times per week)
<sup>b</sup> change in base pairs
<sup>c</sup> CI = 95% confidence interval
<sup>d</sup> Reference category is first quartile or tertile of exposure
\* Statistically significant (p≤0.05)

### Flame retardant exposure and telomere length

We measured 10 flame retardant metabolites in urine, two of which (5-OH-BDE 47 and 5-OH-BDE 100) had no levels above the LOD. Descriptive statistics of flame retardant data revealed disparate distributions of chemical concentrations between occupational groups, with firefighters’ concentrations measured at higher detection frequencies and higher concentrations relative to office workers (**Table 1** and **Additional file 3**). A more in-depth description of the differences in flame retardant concentrations between occupational groups and associated covariates is discussed in Trowbridge et al., 2020 (68). In brief, BDCPP, BCEP, DBuP, and DpCP were all measured at significantly higher levels among firefighters compared to office workers (**Table 1** and **Additional file 3**). Though it had an overall detection frequency of only 29%, DpCP was modeled as a categorical exposure variable because it had a detection frequency of 42% among firefighters. BDCPP was the only flame retardant metabolite with sufficient detection frequency (≥ 70%) to include in models as a continuous variable for both firefighters and office workers. BCEP and DBuP had ≥ 70% detection frequency among firefighters so were included in firefighter models as continuous variables.

**Table 4** and **Additional file 2** show results from stratified linear models controlling for age and logCreatinine. BDCPP concentrations were negatively associated with TL in firefighters and positively associated with TL in office workers. The effect in office workers was statistically significant, with a doubling in BDCPP concentration associated with a 148 (95% CI 22, 274) bp increase in TL. An interaction term for BDCPP and occupation was significant, suggesting that the BDCPP-TL relationship differs significantly by occupation (p-value≤0.01). In models for BCEP, increasing concentration was significantly associated with decreasing TL, with a doubling in BCEP associated with a 99 (95% CI −194, −5) bp decrease in TL.

**Table 4.** Estimated^a^ base pair change in telomere length for a doubling in OPFR metabolite concentration.

|  | **Firefighter $\Delta bp^b$(CI<sup>c</sup>)** | **Office Worker $\Delta bp$(CI)** |
| --- | --- | --- |
| **BDCPP** | -70 (-184,44) | **148 (22,274)\*** |
| **BCEP** | **-99 (-194,-5)\*** | - |
| **DBuP** | 10 (-113,132) | - |

<sup>a</sup> Models adjusted for age (years) and log-transformed creatinine
<sup>b</sup> change in base pairs
<sup>c</sup> CI = 95% confidence interval
\* statistically significant (p≤0.05)

We also ran models with BCEP, DBuP, TBBPA, and DpCP as categorical variables among both firefighters and office workers (**Table 5 and Additional file 2**). All models were stratified due to disparate detection frequencies between groups, which precluded running single models for the full cohort, save for BDCPP. In categorical models, BCEP and TBBPA showed similar patterns of association with TL, with negative effect estimates in firefighters and positive effect estimates in office workers, however, these effect estimates were not statistically significant.

**Table 5.** Estimated^a^ base pair change in telomere length relative to the reference group (<LOD)

|  | **Firefighters $\Delta bp^b$(CI<sup>c</sup>)** | **Office Workers $\Delta bp$(CI)** |
| --- | --- | --- |
| **BCEP** |  |  |
| <LOD/Ref<sup>d</sup> | - | - |
| LOD-50th% | -557 (-1240, 126) | - |
| >50th% | -578 (-1189, 32) | - |
| >LOD | - | 259 (-257, 774) |
| **DBuP** |  |  |
| <LOD/Ref | - | - |
| LOD-50th% | 12 (-699, 724) | - |
| >50th% | 20 (-646, 687) | - |
| >LOD | - | 134 (-427, 694) |
| **TBBPA** |  |  |
| <LOD/Ref | - | - |
| >LOD | -227 (-705, 251) | 349 (-160, 858) |
| **DpCP** |  |  |
| <LOD/Ref | - | - |
| >LOD | -383 (-868, 101) | -57 (-749, 635) |

<sup>a</sup> Models adjusted for age (years) and log-transformed creatinine
<sup>b</sup> change in base pairs
<sup>c</sup> CI = 95% confidence interval
<sup>d</sup> Reference category is <LOD for both firefighters and office workers

## Discussion

This community-based participatory research study examined cross-sectional relationships between PFAS and flame retardant exposures and TL in female firefighters and office workers in San Francisco, CA. To our knowledge, this is the first study to assess the association between chemical exposures and telomere length in female firefighters, and the first study to assess the association between OPFRs and telomere length.

Our analyses of PFAS data revealed statistically significant positive associations between PFOA and PFOS and telomere length among the full cohort, with larger effect estimates among firefighters. Among firefighters, PFOA, PFOS, PFNA, and PFDA were positively associated with TL. Effect estimates among office workers were mostly positive or null across PFAS. These results suggest that exposure to some PFAS, particularly PFOA, PFOS, PFNA, and PFDA, may be associated with telomere lengthening in female firefighters. Prior studies on PFAS exposure and TL is limited and reports mixed results. Huang et al., 2019 examined PFAS and TL in National Health and Nutrition Examination Survey (NHANES) data and reported a strong positive association between PFOS and leukocyte TL in adults and null associations for other PFAS and TL(17). Vriens et al., 2019 found a negative association between PFOA and leukocyte TL in adults aged 50 to 65 years using multipollutant models(18). Zota et al., 2018 similarly used multipollutant models and found no significant associations between prenatal PFAS exposure and repeated measures of leukocyte TL in overweight and obese low-income mothers with an average age of 27.9 years(98). While the literature on the PFAS and TL relationship seems equivocal, such studies may not be comparable due to underlying differences in study populations, methodological approaches, and other confounding and modifying factors.

Our results show that PFAS exposure is associated with telomere lengthening. Previous work has shown that exposure to environmental chemicals is associated with longer TL(16,19,99,100). Mitro et al., 2016 proposed that certain POPs, particularly polychlorinated biphenyls (PCBs), activate the aryl hydrocarbon receptor (AhR), which up-regulates telomerase and may therefore promote cancer(19). Telomerase activation is necessary for cell immortality, which is in turn necessary for tumorigenesis(101). There is some limited evidence of AhR activation by PFAS(102), and so telomerase activation may play an important role in the PFAS-TL relationship. More experimental research that includes the measurement of telomerase is needed to further elucidate potential mechanisms.

Results from flame retardant analyses revealed different effects on TL by occupational status, with flame retardant exposure among firefighters associated with a decrease in TL, and exposure among office workers associated with an increase in TL. However, results were statistically significant only for BDCPP and TL in office workers, and BCEP and TL in firefighters. These differences in effects may not be comparable across occupational groups due to the significantly higher exposure levels and detection frequencies of flame retardant metabolites in firefighters relative to office workers.

Pending further work to characterize the exposure-response relationship, these findings align with other research that has documented variable impacts on TL by dose of environmental chemicals. For instance, Zhang et al., 2003 showed that low doses of arsenite *in vitro* promoted telomerase activity, sustained or lengthened telomeres, and increased cell proliferation, while higher doses of arsenite decreased telomerase and telomere length and promoted apoptosis(103). Similar findings were reported by Ferrario et al., 2009(104). Shin et al., 2010 reported an analogous trend with POPs and TL in NHANEs data, finding longer TL at lower concentrations of POPs and decreased lengthening as POP concentration increased(99).

In both the PFAS-TL and flame retardant-TL analyses, effect estimates differed by occupation. In the PFAS-TL relationship, the differences were in magnitude and estimates of statistical interaction were not significant. In the flame retardant-TL relationship, the differences were in direction and the estimate of statistical interaction between occupation and BDCPP in the BDCPP-TL relationship was significant. While the effect modification by occupation seen in the flame retardant-TL relationship may be attributable to variable effects by dose, it is also possible that there are unmeasured co-exposures affecting TL.

Firefighters are occupationally exposed to many different chemicals including benzene, PAHs, formaldehyde, dioxins, and PBDEs(60–65). Effect estimate differences may be due to unmeasured confounding, including unmeasured chemical co-exposures in firefighters that also have an impact on TL. Non-targeted and/or exposomic approaches are required to improve the characterization of exposures to chemical mixtures and their effects on biological response markers, including TL(105–107).

This was a cross-sectional study, which precludes causal inference (Allen, 2017). Exposure misclassification from cross-sectional sampling may be less relevant when analyzing serum levels of the PFAS assessed here due to their relatively long half-lives in the body(108–110). Flame retardant metabolites were measured in single spot urine samples so temporal variability in concentrations could result in exposure misclassification; however, prior studies indicate that there is temporal stability in OPFR metabolite measurements in urine(111,112). Furthermore, we accounted for urine dilution by including creatinine measurements in our models. Although specific gravity may be considered a preferable measure of urine dilution(113), a study assessing variability in organophosphate metabolite measurements in urine found that temporal variability of creatinine-adjusted metabolite concentrations was lower than that of specific gravity-adjusted and unadjusted metabolite concentrations(111).

## Conclusion

We found positive associations between PFOA and PFOS and telomere length in women workers, with larger effects seen among firefighters compared to office workers for PFOA, PFOS, PFDA, and PFNA. The OPFR metabolites BDCPP and BCEP may also be associated with altered telomere length in women workers. While further exposomic and mechanistic research is needed to more holistically characterize exposures and confirm their relationships with telomere length, the associations reported here suggest mechanisms by which these chemicals may affect carcinogenesis and other adverse health outcomes.

## Data Availability

Data and code may be made available upon request to corresponding author(s).

## Abbreviations

WWBC: Women Workers Biomonitoring Collaborative
TL: telomere length
PFAS: polyfluoroalkyl substances
PFOA: perfluorooctanoic acid
PFOS: perfluorooctane sulfonic acid
PFNA: perfluorononanoic acid
PFDA: perfluorodecanoic acid
PFHxS: perfluorohexane sulfonic acid
PFuNDA: perfluoroundecanoic acid
PFBuS: perfluorobutane sulfonic acid
PFBA: perfluorobutanoic acid
PFHxA: perfluorohexanoic acid
PFDoA: perfluorododecanoic acid
PFHpA: perfluoroheptanoic acid
PFOSA: perfluorooctane sulfonamide
OPFR: organophosphate flame retardant
BDCPP: bis(1,3-dichloro-2-propyl) phosphate
BCEP: bis(2-chloroethyl) phosphate
DbuP: dibutyl phosphate
DBzP: dibenzyl phosphate
DpCP: di-p-cresyl phosphate
DoCP: di-o-cresyl phosphate
TBBA: 2,3,4,5-tetrabromobenzoic acid
TBBPA: tetrabromobisphenol a
PAH: polycyclic aromatic hydrocarbon
PBDE: polybrominated diphenyl ether
LC-MS/MS: liquid chromatography-mass spectrometry
GM: geometric mean
GSD: geometric standard deviation
CI: confidence interval
LOD: limit of detection
DF: detection frequency

## Declarations

### Ethics approval and consent to participate

Informed consent was obtained from all participants prior to data collection activities following protocols approved by the Institutional Review Board of the University of California, Berkeley (#2013-07-5512).

### Consent for publication

Not applicable

### Availability of data and materials

The datasets generated and analyzed for this study are not publicly available because they contain personally identifiable information. They may be made available from the corresponding author on reasonable request.

## Authors’ contributions

CC analyzed and interpreted the data and drafted the manuscript; JT made substantial contributions to study design, acquisition of data, and manuscript revisions; RG, KO, and MM generated chemicals and biomarker data and contributed to manuscript revisions; VB, RAR, HB, and RMF conceived and designed the study, critically discussed results, and revised manuscript drafts. All authors have approved this manuscript.

## Acknowledgements

The authors thank all of the WWBC study participants for their contribution to the study. We thank Anthony Stefani, Emily O’Rourke, Nancy Carmona, Karen Kerr, Julie Mau, Natasha Parks, Lisa Holdcroft, San Francisco Fire Chief Jeanine Nicholson, former San Francisco Fire Chief Joanne Hayes-White, Sharyle Patton, Connie Engel and Nancy Buermeyer for their contributions to the study.

## Authors’ information

RAR and VB, are employed at the Silent Spring Institute, a scientific research organization dedicated to studying environmental factors in women’s health. The Institute is a 501(c)3 public charity funded by federal grants and contracts, foundation grants, and private donations, including from breast cancer organizations. HB is former president and member of United Fire Service Women, a 501(c)3 public charity dedicated to supporting the welfare of women in the San Francisco Fire Department.

## References

1. International Agency for Research on Cancer. IARC Monographs on the Evaluation of Carcinogenic Risks to Humans: Painting, Firefighting, and Shiftwork. [Internet]. Lyon, France: World Health Organization; 2010. Report No.: Vol. 98. Available from: https://monographs.iarc.fr/wp-content/uploads/2018/06/mono98.pdf

2. Daniels RD, Kubale TL, Yiin JH, Dahm MM, Hales TR, Baris D, et al. Mortality and cancer incidence in a pooled cohort of US firefighters from San Francisco, Chicago and Philadelphia (1950–2009). Occup Environ Med. 2014 Jun 1;71(6):388.

3. LeMasters GK, Genaidy AM, Succop P, Deddens J, Sobeih T, Barriera-Viruet H, et al. Cancer Risk Among Firefighters: A Review and Meta-analysis of 32 Studies. J Occup Environ Med [Internet]. 2006;48(11). Available from: https://journals.lww.com/joem/Fulltext/2006/11000/Cancer_Risk_Among_FirefightersA_Review_and.14.aspx

4. Ahn Y-S, Jeong K-S, Kim K-S. Cancer morbidity of professional emergency responders in Korea. Am J Ind Med. 2012 Sep 1;55(9):768–78.

5. Bates MN. Registry-based case–control study of cancer in California firefighters. Am J Ind Med. 2007 May 1;50(5):339–44.

6. Delahunt B, Bethwaite PB, Nacey JN. Occupational risk lor renal cell carcinoma. A case-control study based on the New Zealand Cancer Registry. Br J Urol. 1995 May 1;75(5):578–82.

7. Kang D, Davis LK, Hunt P, Kriebel D. Cancer incidence among male Massachusetts firefighters, 1987–2003. Am J Ind Med. 2008 May 1;51(5):329–35.

8. Ma F, Fleming LE, Lee DJ, Trapido E, Gerace TA, Lai H, et al. Mortality in Florida professional firefighters, 1972 to 1999. Am J Ind Med. 2005 Jun 1;47(6):509–17.

9. Tsai RJ, Luckhaupt SE, Schumacher P, Cress RD, Deapen DM, Calvert GM. Risk of cancer among firefighters in California, 1988–2007. Am J Ind Med. 2015 Jul 1;58(7):715–29.

10. Jalilian H, Ziaei M, Weiderpass E, Rueegg CS, Khosravi Y, Kjaerheim K. Cancer incidence and mortality among firefighters. Int J Cancer. 2019 Nov 15;145(10):2639–46.

11. Evarts B, Stein GP. US Fire Department Profile 2018 [Internet]. National Fire Protection Association; 2020 Feb. Available from: https://www.nfpa.org/-/media/Files/News-and-Research/Fire-statistics-and-reports/Emergency-responders/osfdprofile.pdf

12. Hulett D, Bendick M, Sheila Y, Thomas F, Moccio. Enhancing Women’s Inclusion in Firefighting. Int J Divers Organ Communities Nations. 2007 Nov 1;8.

13. Miller A, Clery S, Richardson S, Topper A, Cronen S, Lilly S, et al. Promising Practices for Increasing Diversity Among First Responders [Internet]. San Francisco: U.S. Department of Labor, Chief Evaluation Office; 2016 Dec. Available from: https://www.dol.gov/sites/dolgov/files/OASP/legacy/files/FirstResponders_SFFDCase_Study.pdf

14. Lee DJ, Koru-Sengul T, Hernandez MN, Caban-Martinez AJ, McClure LA, Mackinnon JA, et al. Cancer risk among career male and female Florida firefighters: Evidence from the Florida Firefighter Cancer Registry (1981-2014). Am J Ind Med. 2020 Apr 1;63(4):285–99.

15. Hou L, Wang S, Dou C, Zhang X, Yu Y, Zheng Y, et al. Air pollution exposure and telomere length in highly exposed subjects in Beijing, China: A repeated-measure study. Environ Int. 2012 Nov 1;48:71–7.

16. Callahan CL, Pavuk M, Birnbaum LS, Ren X, Olson JR, Bonner MR. Serum polychlorinated biphenyls and leukocyte telomere length in a highly-exposed population: The Anniston Community Health Survey. Environ Int. 2017 Nov 1;108:212–20.

17. Huang H, Wang Q, He X, Wu Y, Xu C. Association between polyfluoroalkyl chemical concentrations and leucocyte telomere length in US adults. Sci Total Environ. 2019 Feb 25;653:547–53.

18. Vriens A, Nawrot TS, Janssen BG, Baeyens W, Bruckers L, Covaci A, et al. Exposure to Environmental Pollutants and Their Association with Biomarkers of Aging: A Multipollutant Approach. Environ Sci Technol. 2019 May 21;53(10):5966–76.

19. Mitro SD, Birnbaum LS, Needham BL, Zota AR. Cross-sectional Associations between Exposure to Persistent Organic Pollutants and Leukocyte Telomere Length among U.S. Adults in NHANES, 2001–2002. Environ Health Perspect. 2016;124(5):651–8.

20. Blackburn EH. The molecular structure of centromeres and telomeres. Annu Rev Biochem. 1984;53:163–94.

21. de Lange T. How shelterin solves the telomere end-protection problem. Cold Spring Harb Symp Quant Biol. 2010;75:167–77.

22. Greider CW, Blackburn EH. Identification of a specific telomere terminal transferase activity in Tetrahymena extracts. Cell. 1985 Dec;43(2 Pt 1):405–13.

23. Blackburn EH. Switching and signaling at the telomere. Cell. 2001 Sep 21;106(6):661–73.

24. Blackburn EH, Epel ES, Lin J. Human telomere biology: A contributory and interactive factor in aging, disease risks, and protection. Science. 2015 Dec 4;350(6265):1193–8.

25. Mathon NF, Lloyd AC. Cell senescence and cancer. Nat Rev Cancer. 2001 Dec 1;1(3):203–13.

26. Harley CB, Futcher AB, Greider CW. Telomeres shorten during ageing of human fibroblasts. Nature. 1990 May 1;345(6274):458–60.

27. Blasco MA. Telomere length, stem cells and aging. Nat Chem Biol. 2007 Oct;3(10):640–9.

28. Müezzinler A, Zaineddin AK, Brenner H. A systematic review of leukocyte telomere length and age in adults. Ageing Res Rev. 2013 Mar 1;12(2):509–19.

29. Chen W, Kimura M, Kim S, Cao X, Srinivasan SR, Berenson GS, et al. Longitudinal versus Cross-sectional Evaluations of Leukocyte Telomere Length Dynamics: Age-Dependent Telomere Shortening is the Rule. J Gerontol Ser A. 2011 Jan 3;66A(3):312–9.

30. Cawthon RM, Smith KR, O’Brien E, Sivatchenko A, Kerber RA. Association between telomere length in blood and mortality in people aged 60 years or older. The Lancet. 2003 Feb 1;361(9355):393–5.

31. D’Mello Matthew J.J., Ross Stephanie A., Briel Matthias, Anand Sonia S., Gerstein Hertzel, Paré Guillaume. Association Between Shortened Leukocyte Telomere Length and Cardiometabolic Outcomes. Circ Cardiovasc Genet. 2015 Feb 1;8(1):82–90.

32. Haycock PC, Heydon EE, Kaptoge S, Butterworth AS, Thompson A, Willeit P. Leucocyte telomere length and risk of cardiovascular disease: systematic review and meta-analysis. BMJ. 2014 Jul 8;349:g4227.

33. Zhao J, Miao K, Wang H, Ding H, Wang DW. Association between Telomere Length and Type 2 Diabetes Mellitus: A Meta-Analysis. PLOS ONE. 2013 Nov 21;8(11):e79993.

34. Cohen S, Janicki-Deverts D, Turner RB, Casselbrant ML, Li-Korotky H-S, Epel ES, et al. Association between telomere length and experimentally induced upper respiratory viral infection in healthy adults. JAMA. 2013 Feb 20;309(7):699–705.

35. Wentzensen IM, Mirabello L, Pfeiffer RM, Savage SA. The Association of Telomere Length and Cancer: a Meta-analysis. Cancer Epidemiol Biomark Amp Prev. 2011 Jun 1;20(6):1238.

36. Ma H, Zhou Z, Wei S, Liu Z, Pooley KA, Dunning AM, et al. Shortened Telomere Length Is Associated with Increased Risk of Cancer: A Meta-Analysis. PLOS ONE. 2011 Jun 10;6(6):e20466.

37. Zhang X, Lin S, Funk WE, Hou L. Environmental and occupational exposure to chemicals and telomere length in human studies. Occup Environ Med. 2013 Oct 1;70(10):743.

38. Caini S, Raimondi S, Johansson H, De Giorgi V, Zanna I, Palli D, et al. Telomere length and the risk of cutaneous melanoma and non-melanoma skin cancer: a review of the literature and meta-analysis. J Dermatol Sci. 2015 Dec 1;80(3):168–74.

39. Telomeres Mendelian Randomization Collaboration, Haycock PC, Burgess S, Nounu A, Zheng J, Okoli GN, et al. Association Between Telomere Length and Risk of Cancer and Non-Neoplastic Diseases: A Mendelian Randomization Study. JAMA Oncol. 2017 May 1;3(5):636–51.

40. Pellatt AJ, Wolff RK, Torres-Mejia G, John EM, Herrick JS, Lundgreen A, et al. Telomere length, telomere-related genes, and breast cancer risk: the breast cancer health disparities study. Genes Chromosomes Cancer. 2013/04/30 ed. 2013 Jul;52(7):595–609.

41. Gramatges MM, Telli ML, Balise R, Ford JM. Longer Relative Telomere Length in Blood from Women with Sporadic and Familial Breast Cancer Compared with Healthy Controls. Cancer Epidemiol Biomark Amp Prev. 2010 Feb 1;19(2):605.

42. Qu S, Wen W, Shu X-O, Chow W-H, Xiang Y-B, Wu J, et al. Association of Leukocyte Telomere Length With Breast Cancer Risk: Nested Case-Control Findings From the Shanghai Women’s Health Study. Am J Epidemiol. 2013 Feb 25;177(7):617–24.

43. Samavat H, Xun X, Jin A, Wang R, Koh W-P, Yuan J-M. Association between prediagnostic leukocyte telomere length and breast cancer risk: the Singapore Chinese Health Study. Breast Cancer Res. 2019 Apr 17;21(1):50.

44. Hackett JA, Greider CW. Balancing instability: dual roles for telomerase and telomere dysfunction in tumorigenesis. Oncogene. 2002 Jan 1;21(4):619–26.

45. Hug N, Lingner J. Telomere length homeostasis. Chromosoma. 2006 Dec 1;115(6):413–25.

46. Meena J, Rudolph K, Günes C. Telomere Dysfunction, Chromosomal Instability and Cancer. Recent Results Cancer Res Fortschritte Krebsforsch Prog Dans Rech Sur Cancer. 2015 Sep 17;200:61–79.

47. Adetona O, Zhang J (Junfeng), Hall DB, Wang J-S, Vena JE, Naeher LP. Occupational exposure to woodsmoke and oxidative stress in wildland firefighters. Sci Total Environ. 2013 Apr 1;449:269–75.

48. Fent KW, Eisenberg J, Snawder J, Sammons D, Pleil JD, Stiegel MA, et al. Systemic Exposure to PAHs and Benzene in Firefighters Suppressing Controlled Structure Fires. Ann Occup Hyg. 2014 Jun 6;58(7):830–45.

49. Fent KW, Evans DE, Babik K, Striley C, Bertke S, Kerber S, et al. Airborne contaminants during controlled residential fires. J Occup Environ Hyg. 2018 May 4;15(5):399–412.

50. Navarro KM, Cisneros R, Noth EM, Balmes JR, Hammond SK. Occupational Exposure to Polycyclic Aromatic Hydrocarbon of Wildland Firefighters at Prescribed and Wildland Fires. Environ Sci Technol. 2017 Jun 6;51(11):6461–9.

51. Pleil JD, Stiegel MA, Fent KW. Exploratory breath analyses for assessing toxic dermal exposures of firefighters during suppression of structural burns. J Breath Res. 2014 Sep 4;8(3):037107.

52. Bolstad-Johnson D, Burgess J, Crutchfield C, Storment S, Gerkin R, Wilson J. Characterization of Firefighter Exposures During Fire Overhaul. AIHAJ J Sci Occup Environ Health Saf. 2000 Sep 1;61:636–41.

53. Gainey SJ, Horn GP, Towers AE, Oelschlager ML, Tir VL, Drnevich J, et al. Exposure to a firefighting overhaul environment without respiratory protection increases immune dysregulation and lung disease risk. PLOS ONE. 2018 Aug 21;13(8):e0201830.

54. Alexander BM, Baxter CS. Flame-retardant contamination of firefighter personal protective clothing - A potential health risk for firefighters. J Occup Environ Hyg. 2016 Sep;13(9):D148–155.

55. Brown FR, Whitehead TP, Park J-S, Metayer C, Petreas MX. Levels of non- polybrominated diphenyl ether brominated flame retardants in residential house dust samples and fire station dust samples in California. Environ Res. 2014 Nov 1;135:9–14.

56. Fent KW, Evans DE, Booher D, Pleil JD, Stiegel MA, Horn GP, et al. Volatile Organic Compounds Off-gassing from Firefighters’ Personal Protective Equipment Ensembles after Use. J Occup Environ Hyg. 2015 Jun 3;12(6):404–14.

57. Laitinen JA, Koponen J, Koikkalainen J, Kiviranta H. Firefighters’ exposure to perfluoroalkyl acids and 2-butoxyethanol present in firefighting foams. Adv Biol Monit Occup Environ Health - II. 2014 Dec 1;231(2):227–32.

58. Shen B, Whitehead TP, McNeel S, Brown FR, Dhaliwal J, Das R, et al. High Levels of Polybrominated Diphenyl Ethers in Vacuum Cleaner Dust from California Fire Stations. Environ Sci Technol. 2015 Apr 21;49(8):4988–94.

59. Bott RC, Kirk KM, Logan MB, Reid DA. Diesel particulate matter and polycyclic aromatic hydrocarbons in fire stations. Environ Sci Process Impacts. 2017;19(10):1320–6.

60. Caux C, O’Brien C, Viau C. Determination of Firefighter Exposure to Polycyclic Aromatic Hydrocarbons and Benzene During Fire Fighting Using Measurement of Biological Indicators. Appl Occup Environ Hyg. 2002 May 1;17(5):379–86.

61. Feunekes FDJR, Jongeneelen FJ, Laana H v. d., Schoonhof FHG. Uptake of Polycyclic Aromatic Hydrocarbons Among Trainers in a Fire-Fighting Training Facility. Am Ind Hyg Assoc J. 1997 Jan 1;58(1):23–8.

62. Moen BE, Øvrebø S. Assessment of Exposure to Polycyclic Aromatic Hydrocarbons During Firefighting by Measurement of Urinary 1-Hydroxypyrene. J Occup Environ Med [Internet]. 1997;39(6). Available from: https://journals.lww.com/joem/Fulltext/1997/06000/Assessment_of_Exposure_to_Polycyclic_Aromatic.5.aspx

63. Waldman JM, Gavin Q, Anderson M, Hoover S, Alvaran J, Ip HSS, et al. Exposures to environmental phenols in Southern California firefighters and findings of elevated urinary benzophenone-3 levels. Environ Int. 2016 Mar;88:281–7.

64. Dobraca D, Israel L, McNeel S, Voss R, Wang M, Gajek R, et al. Biomonitoring in California firefighters: metals and perfluorinated chemicals. J Occup Environ Med. 2015 Jan;57(1):88–97.

65. Shaw SD, Berger ML, Harris JH, Yun SH, Wu Q, Liao C, et al. Persistent organic pollutants including polychlorinated and polybrominated dibenzo-p-dioxins and dibenzofurans in firefighters from Northern California. Chemosphere. 2013 Jun 1;91(10):1386–94.

66. Fent KW, LaGuardia M, Luellen D, McCormick S, Mayer A, Chen I-C, et al. Flame retardants, dioxins, and furans in air and on firefighters’ protective ensembles during controlled residential firefighting. Environ Int. 2020 Jul 1;140:105756.

67. Trowbridge J, Gerona RR, Lin T, Rudel RA, Bessonneau V, Buren H, et al. Exposure to Perfluoroalkyl Substances in a Cohort of Women Firefighters and Office Workers in San Francisco. Environ Sci Technol. 2020 Mar 17;54(6):3363–74.

68. Trowbridge J, Gerona R, McMaster M, Ona K, Clarity C, Bessonneau V, et al. Organophosphate and organohalogen flame-retardant exposure and thyroid hormone disruption in a cohort of female firefighters and office workers from San Francisco. medRxiv. 2020 Jan 1;2020.10.06.20207498.

69. Agency for Toxic Substances and Disease Registry. Toxicological Profile for Perfluoroalkyls [Internet]. 2018 Jun. Available from: https://www.atsdr.cdc.gov/toxprofiles/tp200.pdf

70. Moody CA, Field JA. Perfluorinated Surfactants and the Environmental Implications of Their Use in Fire-Fighting Foams. Environ Sci Technol. 2000 Sep 1;34(18):3864–70.

71. Jin C, Sun Y, Islam A, Qian Y, Ducatman A. Perfluoroalkyl Acids Including Perfluorooctane Sulfonate and Perfluorohexane Sulfonate in Firefighters. J Occup Environ Med [Internet]. 2011;53(3). Available from: https://journals.lww.com/joem/Fulltext/2011/03000/Perfluoroalkyl_Acids_Including_Perfluorooctane.17.aspx

72. Barry Vaughn, Winquist Andrea, Steenland Kyle. Perfluorooctanoic Acid (PFOA) Exposures and Incident Cancers among Adults Living Near a Chemical Plant. Environ Health Perspect. 2013 Jan 1;121(11–12):1313–8.

73. Lopez-Espinosa Maria-Jose, Mondal Debapriya, Armstrong Ben, Bloom Michael S., Fletcher Tony. Thyroid Function and Perfluoroalkyl Acids in Children Living Near a Chemical Plant. Environ Health Perspect. 2012 Jul 1;120(7):1036–41.

74. Lopez-Espinosa Maria-Jose, Mondal Debapriya, Armstrong Ben G., Eskenazi Brenda, Fletcher Tony. Perfluoroalkyl Substances, Sex Hormones, and Insulin-like Growth Factor-1 at 6–9 Years of Age: A Cross-Sectional Analysis within the C8 Health Project. Environ Health Perspect. 2016 Aug 1;124(8):1269–75.

75. Liu H, Chen Q, Lei L, Zhou W, Huang L, Zhang J, et al. Prenatal exposure to perfluoroalkyl and polyfluoroalkyl substances affects leukocyte telomere length in female newborns. Environ Pollut. 2018 Apr 1;235:446–52.

76. Steenland Kyle, Zhao Liping, Winquist Andrea, Parks Christine. Ulcerative Colitis and Perfluorooctanoic Acid (PFOA) in a Highly Exposed Population of Community Residents and Workers in the Mid-Ohio Valley. Environ Health Perspect. 2013 Aug 1;121(8):900–5.

77. Joensen Ulla Nordström, Bossi Rossana, Leffers Henrik, Jensen Allan Astrup, Skakkebæk Niels E., Jørgensen Niels. Do Perfluoroalkyl Compounds Impair Human Semen Quality? Environ Health Perspect. 2009 Jun 1;117(6):923–7.

78. Bassler J, Ducatman A, Elliott M, Wen S, Wahlang B, Barnett J, et al. Environmental perfluoroalkyl acid exposures are associated with liver disease characterized by apoptosis and altered serum adipocytokines. Environ Pollut. 2019 Apr 1;247:1055–63.

79. Blake BE, Pinney SM, Hines EP, Fenton SE, Ferguson KK. Associations between longitudinal serum perfluoroalkyl substance (PFAS) levels and measures of thyroid hormone, kidney function, and body mass index in the Fernald Community Cohort. Environ Pollut. 2018 Nov 1;242:894–904.

80. Park J-S, Voss RW, McNeel S, Wu N, Guo T, Wang Y, et al. High exposure of California firefighters to polybrominated diphenyl ethers. Environ Sci Technol. 2015 Mar 3;49(5):2948–58.

81. US Environmental Protection Agency. Polybrominated Diphenyl Ethers (PBDEs) Action Plan [Internet]. 2009 Dec. Available from: https://www.epa.gov/sites/production/files/2015-09/documents/pbdes_ap_2009_1230_final.pdf

82. Stapleton HM, Sharma S, Getzinger G, Ferguson PL, Gabriel M, Webster TF, et al. Novel and High Volume Use Flame Retardants in US Couches Reflective of the 2005 PentaBDE Phase Out. Environ Sci Technol. 2012 Dec 18;46(24):13432–9.

83. Dodson RE, Perovich LJ, Covaci A, Van den Eede N, Ionas AC, Dirtu AC, et al. After the PBDE Phase-Out: A Broad Suite of Flame Retardants in Repeat House Dust Samples from California. Environ Sci Technol. 2012 Dec 18;46(24):13056–66.

84. Shen B, Whitehead TP, Gill R, Dhaliwal J, Brown FR, Petreas M, et al. Organophosphate flame retardants in dust collected from United States fire stations. Environ Int. 2018 Mar 1;112:41–8.

85. Meeker John D., Stapleton Heather M. House Dust Concentrations of Organophosphate Flame Retardants in Relation to Hormone Levels and Semen Quality Parameters. Environ Health Perspect. 2010 Mar 1;118(3):318–23.

86. Preston EV, McClean MD, Claus Henn B, Stapleton HM, Braverman LE, Pearce EN, et al. Associations between urinary diphenyl phosphate and thyroid function. Environ Int. 2017 Apr 1;101:158–64.

87. Liu X, Ji K, Choi K. Endocrine disruption potentials of organophosphate flame retardants and related mechanisms in H295R and MVLN cell lines and in zebrafish. Aquat Toxicol.2012 Jun 15;114–115:173–81.

88. Xu T, Wang Q, Shi Q, Fang Q, Guo Y, Zhou B. Bioconcentration, metabolism and alterations of thyroid hormones of Tris(1,3-dichloro-2-propyl) phosphate (TDCPP) in Zebrafish. Environ Toxicol Pharmacol. 2015 Sep 1;40(2):581–6.

89. Wang Q, Lai NL-S, Wang X, Guo Y, Lam PK-S, Lam JC-W, et al. Bioconcentration and Transfer of the Organophorous Flame Retardant 1,3-Dichloro-2-propyl Phosphate Causes Thyroid Endocrine Disruption and Developmental Neurotoxicity in Zebrafish Larvae. Environ Sci Technol. 2015 Apr 21;49(8):5123–32.

90. Oliveri AN, Bailey JM, Levin ED. Developmental exposure to organophosphate flame retardants causes behavioral effects in larval and adult zebrafish. Investig Neurotox Past Present Future Flame Retard. 2015 Nov 1;52:220–7.

91. Chen G, Jin Y, Wu Y, Liu L, Fu Z. Exposure of male mice to two kinds of organophosphate flame retardants (OPFRs) induced oxidative stress and endocrine disruption. Environ Toxicol Pharmacol. 2015 Jul 1;40(1):310–8.

92. Møller P, Wils RS, Jensen DM, Andersen MHG, Roursgaard M. Telomere dynamics and cellular senescence: an emerging field in environmental and occupational toxicology. Crit Rev Toxicol. 2018 Oct 21;48(9):761–88.

93. Ock J, Kim J, Choi Y-H. Organophosphate insecticide exposure and telomere length in U.S. adults. Sci Total Environ. 2020 Mar 20;709:135990.

94. Benjamini Y, Hochberg Y. Controlling the False Discovery Rate: A Practical and Powerful Approach to Multiple Testing. J R Stat Soc Ser B Methodol. 1995;57(1):289–300.

95. RStudio Team. RStudio: Integrated Development for R [Internet]. Boston, MA: RStudio, Inc.; 2016. Available from: http://www.rstudio.com/

96. R Development Core Team. R: A Language and Environment for Statistical Computing [Internet]. Vienna, Austria: R Foundation for Statitical Computing; 2018. Available from: https://www.R-project.org/

97. Barr Dana B., Wilder Lynn C., Caudill Samuel P., Gonzalez Amanda J., Needham Lance L., Pirkle James L. Urinary Creatinine Concentrations in the U.S. Population: Implications for Urinary Biologic Monitoring Measurements. Environ Health Perspect. 2005 Feb 1;113(2):192–200.

98. Zota AR, Geller RJ, Romano LE, Coleman-Phox K, Adler NE, Parry E, et al. Association between persistent endocrine-disrupting chemicals (PBDEs, OH-PBDEs, PCBs, and PFASs) and biomarkers of inflammation and cellular aging during pregnancy and postpartum. Environ Int. 2018 Jun 1;115:9–20.

99. Shin J-Y, Choi YY, Jeon H-S, Hwang J-H, Kim S-A, Kang J-H, et al. Low-dose persistent organic pollutants increased telomere length in peripheral leukocytes of healthy Koreans. Mutagenesis. 2010 Jul 8;25(5):511–6.

100. Scinicariello F, Buser MC. Polychlorinated Biphenyls and Leukocyte Telomere Length: An Analysis of NHANES 1999–2002. EBioMedicine. 2015 Dec 1;2(12):1974–9.

101. Cong Y-S, Wright WE, Shay JW. Human Telomerase and Its Regulation. Microbiol Mol Biol Rev. 2002 Sep 1;66(3):407.

102. Long M, Ghisari M, Bonefeld-Jørgensen EC. Effects of perfluoroalkyl acids on the function of the thyroid hormone and the aryl hydrocarbon receptor. Environ Sci Pollut Res. 2013 Nov 1;20(11):8045–56.

103. Zhang T-C, Schmitt MT, Mumford JL. Effects of arsenic on telomerase and telomeres in relation to cell proliferation and apoptosis in human keratinocytes and leukemia cells in vitro. Carcinogenesis. 2003 Nov 1;24(11):1811–7.

104. Ferrario D, Collotta A, Carfi M, Bowe G, Vahter M, Hartung T, et al. Arsenic induces telomerase expression and maintains telomere length in human cord blood cells. Toxicology. 2009 Jun 16;260(1):132–41.

105. Rappaport SM. Implications of the exposome for exposure science. J Expo Sci Environ Epidemiol. 2011 Feb;21(1):5–9.

106. Wild CP. Complementing the Genome with an “Exposome”: The Outstanding Challenge of Environmental Exposure Measurement in Molecular Epidemiology. Cancer Epidemiol Biomark Amp Prev. 2005 Aug 1;14(8):1847.

107. Bessonneau V, Rudel RA. Mapping the Human Exposome to Uncover the Causes of Breast Cancer. Int J Environ Res Public Health. 2019 Dec 27;17(1):189.

108. Li Y, Fletcher T, Mucs D, Scott K, Lindh CH, Tallving P, et al. Half-lives of PFOS, PFHxS and PFOA after end of exposure to contaminated drinking water. Occup Environ Med. 2018 Jan 1;75(1):46.

109. Zhang Y, Beesoon S, Zhu L, Martin JW. Biomonitoring of Perfluoroalkyl Acids in Human Urine and Estimates of Biological Half-Life. Environ Sci Technol. 2013 Sep 17;47(18):10619–27.

110. Wu XM, Bennett DH, Calafat AM, Kato K, Strynar M, Andersen E, et al. Serum concentrations of perfluorinated compounds (PFC) among selected populations of children and adults in California. Environ Res. 2014/11/20 ed. 2015 Jan;136:264–73.

111. Wang Y, Li W, Martínez-Moral MP, Sun H, Kannan K. Metabolites of organophosphate esters in urine from the United States: Concentrations, temporal variability, and exposure assessment. Environ Int. 2019 Jan 1;122:213–21.

112. Meeker JD, Cooper EM, Stapleton HM, Hauser R. Urinary metabolites of organophosphate flame retardants: temporal variability and correlations with house dust concentrations. Environ Health Perspect. 2013/03/05 ed. 2013 May;121(5):580–5.

113. Sauvé J-F, Lévesque M, Huard M, Drolet D, Lavoué J, Tardif R, et al. Creatinine and Specific Gravity Normalization in Biological Monitoring of Occupational Exposures. J Occup Environ Hyg. 2015 Feb 1;12(2):123–9.

