## Additional file 1 for "Associations between polyfluoroalkyl substance and organophosphate flame retardant exposures and telomere length in a cohort of women firefighters and office workers in San Francisco" (from another version of this preprint: version 1, 2020-11-07)

**Effect estimates[b] for PFAS exposure on telomere length by occupation from minimally adjusted and fully adjusted[a] continuous linear models.**

| | **Full cohort** (N=163) | | **Firefighters** (N=84) | | **Office Workers** (N=79) | |
|---|---|---|---|---|---|---|
| | **β(CI)[c]** | **p-value** | **β(CI)** | **p-value** | **β(CI)** | **p-value** |
| **PFHxS** | | | | | | |
| Model 1 | 0.15(-0.02, 0.32) | 0.09 | 0.10(-0.13, 0.34) | 0.39 | 0.13(-0.14, 0.40) | 0.35 |
| Model 2 | 0.12(-0.05, 0.29) | 0.17 | 0.13(-0.10, 0.36) | 0.26 | 0.10(-0.17, 0.37) | 0.46 |
| **PFOA** | | | | | | |
| Model 1 | 0.39(0.08, 0.71) | 0.02* | 0.57(0.12, 1.02) | 0.01* | 0.25(-0.19, 0.70) | 0.26 |
| Model 2 | 0.35(0.04, 0.66) | 0.03* | 0.47(0.02, 0.93) | 0.04* | 0.24(-0.20, 0.68) | 0.28 |
| **PFOS** | | | | | | |
| Model 1 | 0.26(0.02, 0.51) | 0.03* | 0.44(0.05, 0.83) | 0.03* | 0.13(-0.18, 0.44) | 0.40 |
| Model 2 | 0.25(0.01, 0.49) | 0.04* | 0.39(-0.01, 0.79) | 0.05* | 0.18(-0.14, 0.49) | 0.26 |
| **PFNA** | | | | | | |
| Model 1 | 0.18(-0.09, 0.45) | 0.18 | 0.15(-0.21, 0.50) | 0.41 | 0.09(-0.34, 0.52) | 0.67 |
| Model 2 | 0.10(-0.17, 0.37) | 0.47 | 0.07(-0.28, 0.43) | 0.68 | 0.17(-0.26, 0.60) | 0.44 |
| **PFDA** | | | | | | |
| Model 1 | 0.20(-0.04, 0.44) | 0.10 | 0.43(0.02, 0.84) | 0.04* | 0.05(-0.25, 0.34) | 0.74 |
| Model 2 | 0.15(-0.08, 0.38) | 0.21 | 0.37(-0.05, 0.78) | 0.08 | 0.09(-0.21, 0.39) | 0.57 |
| **PFUnDA** | | | | | | |
| Model 1 | 0.04(-0.08, 0.15) | 0.53 | 0.11(-0.07, 0.30) | 0.23 | -0.04(-0.20, 0.11) | 0.56 |
| Model 2 | 0.02(-0.10, 0.13) | 0.76 | 0.09(-0.10, 0.27) | 0.35 | 0.00(-0.16, 0.17) | 0.98 |
| **PFBuS** | | | | | | |
| Model 1 | 0.01(-0.10, 0.13) | 0.81 | -0.05(-0.22, 0.12) | 0.57 | 0.07(-0.10, 0.24) | 0.39 |
| Model 2 | 0.03(-0.08, 0.15) | 0.57 | -0.01(-0.18, 0.16) | 0.94 | 0.07(-0.10, 0.24) | 0.44 |

[a] Model 1 adjusted for age (years); Model 2 adjusted for age (years), dairy and egg consumption (times per week), and occupation (in full group only)

[b] Effect estimates indicate change in mean telomere length (kbp) for every log-unit change in PFAS concentration

[c] CI = 95% confidence interval
