## Additional file 2 for "Associations between polyfluoroalkyl substance and organophosphate flame retardant exposures and telomere length in a cohort of women firefighters and office workers in San Francisco" (from another version of this preprint: version 1, 2020-11-07)

**Effect estimates for OPFR exposure on telomere length by occupation from adjusted[a] linear models.**

| | **Firefighters** (N=84) | | **Office workers** (N=79) | |
|---|---|---|---|---|
| | **βcoeff(CI)[c]** | **p-value** | **βcoeff(CI)[c]** | **p-value** |
| **BDCPP** | | | | |
| Continuous Model | -0.10(-0.27, 0.06) | 0.23 | 0.21(0.03, 0.40) | 0.02* |
| **BCEP** | | | | |
| Continuous Model | -0.14(-0.28, -0.01) | 0.04* | - | - |
| Categorical Models | | | | |
| <LOD/Ref[b] | - | - | - | - |
| LOD-50th% | -0.56(-1.24, 0.13) | 0.11 | - | - |
| >50th% | -0.58(-1.19, 0.03) | 0.06 | - | - |
| ≥LOD | - | - | 0.26(-0.26, 0.77) | 0.32 |
| **DBuP** | | | | |
| Continuous Model | 0.01(-0.16, 0.19) | 0.88 | - | - |
| Categorical Models | | | | |
| <LOD/Ref[b] | - | - | - | - |
| LOD-50th% | 0.01(-0.70, 0.72) | 0.97 | - | - |
| >50th% | 0.02(-0.65, 0.69) | 0.95 | - | - |
| ≥LOD | - | - | 0.13(-0.43, 0.69) | 0.64 |
| **TBBPA** | | | | |
| Categorical Model | | | | |
| <LOD/Ref[b] | - | - | - | - |
| ≥LOD | -0.23(-0.70, 0.25) | 0.35 | 0.35(-0.16, 0.86) | 0.18 |
| **DpCP** | | | | |
| Categorical Model | | | | |
| <LOD/Ref[b] | - | - | - | - |
| ≥LOD | -0.38(-0.87, 0.10) | 0.12 | -0.06(-0.75, 0.64) | 0.87 |

[a] Models adjusted for age (years) and log-transformed creatinine

[b] Reference category is <LOD for both firefighters and office workers

[c] CI = 95% confidence interval
