## Additional file 3 for "Associations between polyfluoroalkyl substance and organophosphate flame retardant exposures and telomere length in a cohort of women firefighters and office workers in San Francisco" (from another version of this preprint: version 1, 2020-11-07)

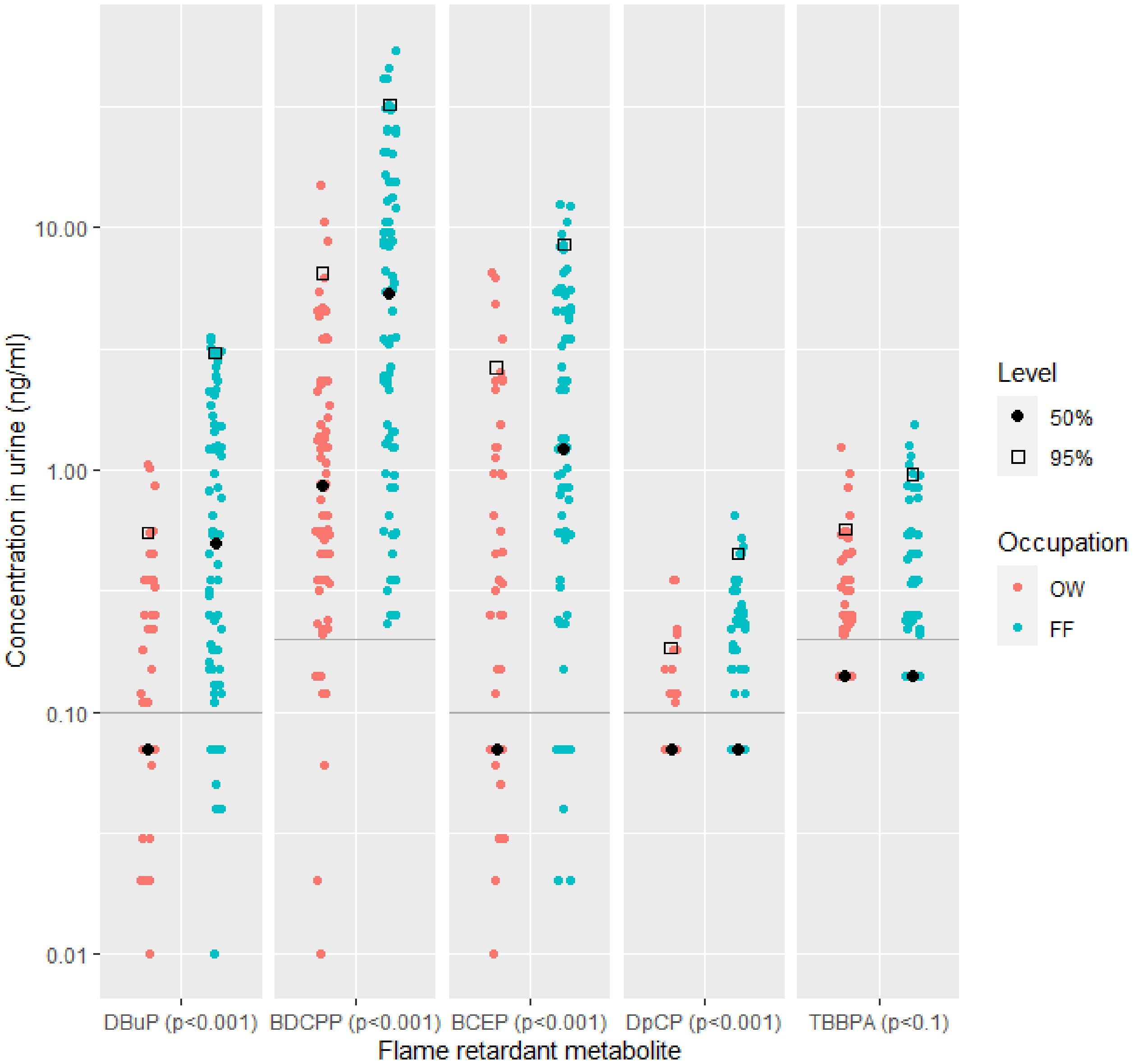


**Flame retardant metabolite concentrations (ng/mL) by occupation**; values below the LOD were substituted with LC-MS/MS reported values below the LOD where available, and LOD/√2 for remaining non-detect values; grey lines represent chemical-specific LODs; p-values indicate significance of permutation tests assessing difference in concentration distributions between occupational groups.
